# Secondary findings in a large Pakistani cohort tested with whole genome sequencing

**DOI:** 10.1101/2022.08.14.22278472

**Authors:** Aliaksandr Skrahin, Huma Arshad Cheema, Maqbool Hussain, Nuzhat Noureen Rana, Khalil Ur Rehman, Raman Kumar, Gabriela Oprea, Najim Ameziane, Arndt Rolfs, Volha Skrahina

**Affiliations:** Arcensus GmbH, Goethestr. 20, 18055 Rostock, Germany; University of Child Health Sciences, the Children’s Hospital, Lahore, Pakistan; Pakistan Institute of Medical Sciences, Islamabad, Pakistan; The Children’s Hospital and the Institute of Child Health, Multan, Pakistan; Town Women and Children Hospital, Peshawar, Pakistan; Liaquat National Hospital, Karachi, Pakistan; University of Rostock, Medical Faculty, Schillingallee 70, 18055 Rostock, Germany

## Abstract

**Background:** Studies in the field of genomic secondary findings (SF) are diverse regarding participants’ characteristics; sequencing methods; versions of the ACMG SF gene list.

**Aim and methods:** Based on whole genome sequencing (WGS) and version 3.1 of ACMG SF list (ACMG SF), we studied SF in 863 individuals from Pakistan: 62% males; 80% had consanguineous parents. In addition to the ACMG SF we have generated a list of gene-disease pairs that have a clear epidemiological and medically actionable value (non-ACMG SF) in Pakistan.

**Results:** The total rate of SF was 4.6%, with rates of ACMG SF – 2.7% and non-AGMG SF – 1.9%. 75.0% of ACMG SF were related to cardiovascular diseases (CVD); cancer predisposition syndromes accounted for 16.7%. Among non-ACMG SF 18.8% belong to eye diseases, followed by neuromuscular – 12.5%, metabolic – 12.5%, and urinary system diseases – 12.5%; CVD accounted for 6.3%. We found high proportion of biallelic mutations among both ACMG (4.2%) and non-ACMG (50%) SF.

**Conclusions:** The frequency of ACMG SF is within the range reported in most studies. High proportion of CVD can be explained by inclusion of additional CVD in the ACMG v3.1 SF list. 1.9% of non-ACMG SF and high proportion of biallelic variants are relevant to epidemiology of Pakistan as a country with high rate of consanguineous marriages. In such countries the ACMG criteria for SF can be expanded, and our list of non-ACMG SF is one example. Our findings may help guide the development of standards of practice in genomic medicine and drive future research.

## INTRODUCTION

The term “incidental finding” is used in clinical molecular diagnostics to refer to the detection of pathogenic or likely pathogenic variants in genes unrelated to the disease for which the sequencing test was intended. In 2013, the American College of Medical Genetics and Genomics (ACMG) published recommendations for reporting incidental findings in clinical sequencing for a list of 56 genes (v1.0) associated with Mendelian disorders that are medically actionable (Green et al. 2013). These genes were selected based on clinical evidence for high penetrance of a severe disease. The list included genes associated with diseases for which preventive measures and/or treatments are available, as well as genes related to diseases for which individuals with a pathogenic variant may be asymptomatic at the time of test request. In addition, evidence-based guidelines were recommended by ACMG and Association for Molecular Pathology (AMP) in 2015 to standardize the clinical interpretation of sequence variants (Richards et al. 2015). The gene list was increased to 59 (v2.0) (Kalia et al. 2017), 73 genes (v3.0) (Miller et al. 2021a), and currently includes 78 genes (v3.1) (Miller et al. 2022).

Furthermore, the term “incidental findings” was replaced by the more meaningful term “secondary findings” (SF). A new framework to update the ACMG SF list annually was also outlined. Studies in the secondary findings area are diverse in design, including the number of participants, their characteristics such as consanguinity, ethnicity, symptoms; the of use of different sequencing methods, whole genome (WGS) or exome sequencing (WES); the use of ACMG SF gene list version (v1.0 SF, 2013; v2.0 SF, 2016; v3.0 SF, 2021); additional analyses of SF not included in the ACMG SF lists.

One of the main problems in applying the ACMG SF guidelines is that, to date, expert opinion on the inclusion of these gene-disease pairs in the ACMG SF list is not based on testing of broader patient populations.

Countries and regions that have specific characteristics in the epidemiology of genetic diseases may have criteria for SF that differ from those recommended by ACMG. In clinical practice and research, based on high frequency of the genetic disorders in the country due to the high level of consanguineous marriages the list of secondary genetic findings can be expanded using the opinions of experts familiar with the regional situation and other relevant resources, e.g., ClinGen and eMERGE. ClinGen (https://www.clinicalgenome.org/) is a National Institutes of Health (NIH)-funded resource dedicated to building an authoritative central resource that defines the clinical relevance of genes and variants for use in precision medicine and research resulted in evidence-based reports and semi-quantitative metric scores for 252 clinically actionable genes. eMERGE (https://emerge-network.org/) is a network organized by the National Human Genome Research Institute (NHGRI) that combines DNA biorepositories with electronic medical record (EMR) systems for large scale, high-throughput genetic research in support of implementing genomic medicine. 109 genes and 1,551 single nucleotide variants (SNV) that allow for clinically actionable, pathogenic variants to be returned were identified and validated by the eMERGEseq platform.

Pakistan is a developing country with an estimated population of over 220 million and a high burden of non-communicable diseases. The rate of consanguineous marriages in Pakistan is estimated to be 46-98%. This is associated with high rates of genetic inherited diseases and infant mortality. (Riaz et al. 2019).

We investigated the prevalence of ACMG v3.1 SF (ACMG SF) in probands who have been subjected to genetic testing by WGS.

In addition to the ACMG SF, we have analysed a cohort of about 1000 Pakistani participants and identified gene-disease pairs with strong medical actionability, not yet included in the ACMG v3.1 secondary finding list. Based on this, we generated a list of Pakistani non-ACMG secondary findings (non-ACMD SF) consisting of 37 genes associated with 39 diseases. (Supplement Table S3).

Concomitantly, genetic variants in ACMG SF and non-ACMG SF lists were identified and categorized as primary findings (ACMG PF and non-ACMG PF, respectively) as they were related to the clinical symptomatology or primary reason for genomic testing.

## RESULTS

### Cohort description

The study cohort consists of 863 Pakistani individuals: 700 (81.1%) index participants and 163 (19.9%) their family members; males 532 (61.7%); most participants, 741 (85.9%) were clinically affected and had various symptoms, their age was 4.9+/-6.2 (mean+/-SD), age of 122 (14.1%) healthy/asymptomatic participants was 28.3+/-10.5 years; 559 (79.9%) participants had consanguineous parents; of those with known self-declared ethnicity majority belong to Punjabi. (Table 1).

**Table 1:** Pakistani cohort characterization.

| **Analysed individuals (n = 863)** | **Total counts** | **%** |
| --- | --- | --- |
| Index patients | 700 | 81.11% |
| Relatives | 163 | 18.89% |
| **Sex (n=863)** | **Total counts** | **%** |
| Males | 532 | 61.65% |
| Females | 331 | 38.35% |
| **Clinical status (n=863)** | **Total counts** | **%** |
| Clinically affected | 741 | 85.86% |
| Healthy/ asymptomatic | 122 | 14.14% |
| **Consanguinity status (n= 700 index patients/families)** | **Total counts** | **%** |
| Consanguineous parents | 559 | 79.86% |
| Non-consanguineous parents | 122 | 17.43% |
| Not disclosed | 19 | 2.71% |
| **Age at genetic testing (n = 863); expressed as years, months** | **Mean** | **SD** |
| Clinically affected (n=741) | 4.93 | 6.17 |
| Healthy/ asymptomatic (n=122) | 28.34 | 10.52 |

### Secondary findings (SF)

#### Secondary findings of the ACMG v3.0 (2021) secondary findings list (ACMG SF)

Twenty-four ACMG SF were detected in 23/863 (2.7%) participants. One participant had two ACMG SF (in *MYBPC3* and *TTN* genes). Two members of one family had the same variant in *TMEM43*. Two unrelated participants have the same variant in *KCNQ1*. There were 23 monoallelic (heterozygous) and 1 (4.2%) biallelic (homozygous) mutations. In total, there were 22 unique variants in 14 unique genes associated with 15 genetic diseases. Only one ACMG SF was found in asymptomatic participant (in *RYR1* gene), giving a frequency 0.8%, while frequency of ACMG SF in symptomatic participants was 3.0% (OR 3.6 [95 % CI: 0.5 to 27.1], p= 0.21). (Supplement Table S1).

18/24 (75.0%) ACMG SF were related to cardiovascular diseases (CVD), among them: *TTN* dilated cardiomyopathy (DCM) - 4; *ACTC1* DCM (1); *MYBPC3* hypertrophic cardiomyopathy (HCM) - 2, *TNNT2* DCM/HCM - 1; *KCNQ1* long QT syndrome (LQTS) - 3; *LDLR* familial hypercholesterolemia (FH) - 2; *PCSK9* FH - 1; *APOB* FH - 1; *TMEM43* arrhythmogenic right ventricular dysplasia (ARVD) - 2; *FBN1* Marfan syndrome - 1. Cancer predisposition syndromes accounted for 16.7% (4/24) of ACMG SF: *MSH6* Lynch syndrome - 1, *BRCA1* hereditary breast and ovarian cancer (HBOC) - 1 and *PALB2* hereditary breast cancer (HBC) - 2. *RYR1* malignant hyperthermia was detected in two (8.3%) participants. (Figure 1).

**Figure 1.**
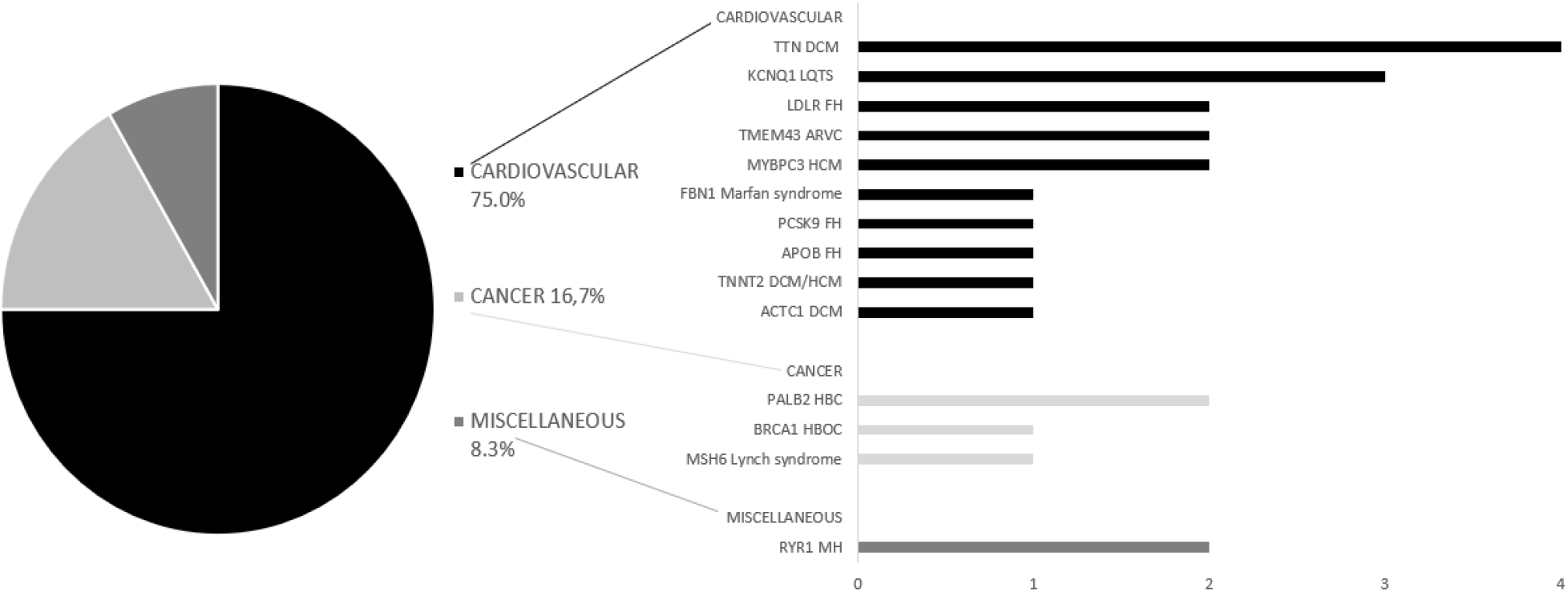
The ACMG v3.0 secondary findings. DCM – dilated cardiomyopathy; LQTS – long QT syndrome; FH – familial hypercholesterolemia; ARVC – arrhythmogenic right ventricular cardiomyopathy; HBC – hereditary breast cancer; HBOC – hereditary breast and ovarian cancer; MH – malignant hyperthermia.

We detected one participant (0.12%) with a heterozygous likely pathogenic (LP) variant in *MUTHYH* gene meaning a carrier status for autosomal recessive (AR) *MUTYH*-related familial adenomatous polyposis.

#### Secondary findings of non-ACMG secondary findings list (non-ACMG SF)

Sixteen non-ACMG SF were detected in 16/863 (1.9%) participants. There were 8 monoallelic (50%) (heterozygous and hemizygous) and 8 (50%) biallelic (homozygous and compound heterozygous) mutations. Totally, there were 17 unique variants in 14 unique genes related to 15 genetic diseases (Supplement Table S1). Among non-ACMG SF 18.8% belong to eye diseases, followed by neuromuscular (12.5%), metabolic (12.5%), and urinary system diseases (12.5%); cardiovascular diseases accounted for 6.3%. (Figure 2). We detect non-ACMG SF in 15/741 (2.0%) symptomatic and 1/122 (0.8%) asymptomatic participants, OR 0.40 (95 % CI: 0.05 – 3.09, p = 0.38). (Supplement Table S1).

**Figure 2.**
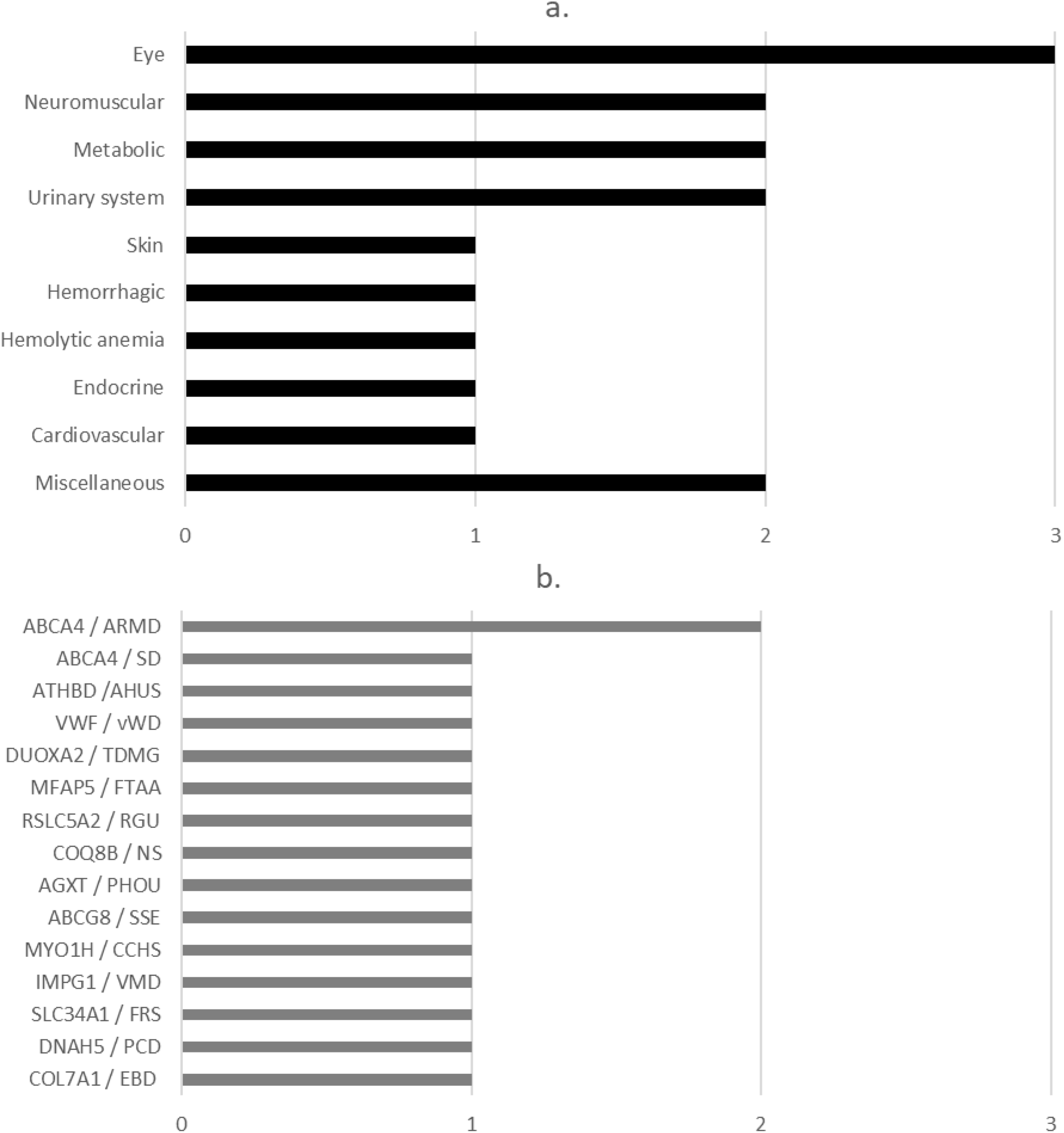
Non-ACMG secondary findings: a. Disease types. B. Genes / Diseases. ARMD - Age-related macular degeneration type 2; SD - Stargardt disease type 1; PCD - Primary ciliary dyskinesia type 3; FRS - Fanconi renotubular syndrome type 2; VMD - Vitelliform macular dystrophy type 4; CCHS - Congenital central hypoventilation syndrome type 2; SSE - Sitosterolemia type 1; PHOU - Primary hyperoxaluria type 1; NS - Nephrotic syndrome; type 9; RGU - Renal glucosuria; FTAA - Familial thoracic aortic aneurysm type 9; TDMG - Thyroid dyshormonogenesis type 5; vWD - von Willebrand disease type 1; AHUS - Atypical hemolytic uremic syndrome type 6; EBD - Epidermolysis bullosa dystrophica.

Eight participant (0.93%) had a carrier status, heterozygous P/LP variants for AR diseases: *CFTR* Cystic fibrosis (5), *ITGB4* Junctional epidermolysis bullosa type 5A (1), *MEFV* AR familial Mediterranean fever (1), and *SLC7A9* AR cystinuria (1). (Supplement Table S1).

### Primary findings (PF)

According to the phenotype and family history of the participants some gene-disease pairs included in both the ACMG SF and non-ACMG SF lists were reported as primary findings (ACMG PF and non-ACMG PF, respectively).

#### Primary findings of the ACMG v3.0 (2021) secondary findings list (ACMG PF)

The ACMG PF, were detected in 41/741 (5.5%) symptomatic participants. For comparison to the ACMG SF, we excluded those with variants of unknown significance (VUS). The ACMG PF that included only P/LP variants were detected in 35/741 (4.7%) symptomatic participants. The same homozygous variants were found among two family members in 3 families: (1) and (2) in BTD gene; and (3) in ATP7B gene. There were 4 (11.4%) monoallelic (heterozygous) and 31 (88.6%) biallelic (homozygous and compound heterozygous) mutations among the ACMG PF. Thus, in contrast to the ACMG SF, biallelic variants predominated among the ACMG PF. In total, there were 23 unique P/LP variants in 8 genes related to 8 genetic diseases (Supplement Table S2).

Most gene-disease pairs belong to either only the ACMG SF or only the ACMG PF. Two gene-disease pairs occurred in both groups: Marfan syndrome (*FBN1*), and familial hypercholesterolemia type 1 (*LDLR*). (Figure 3). No identical variants have been found in both groups.

**Figure 3.**
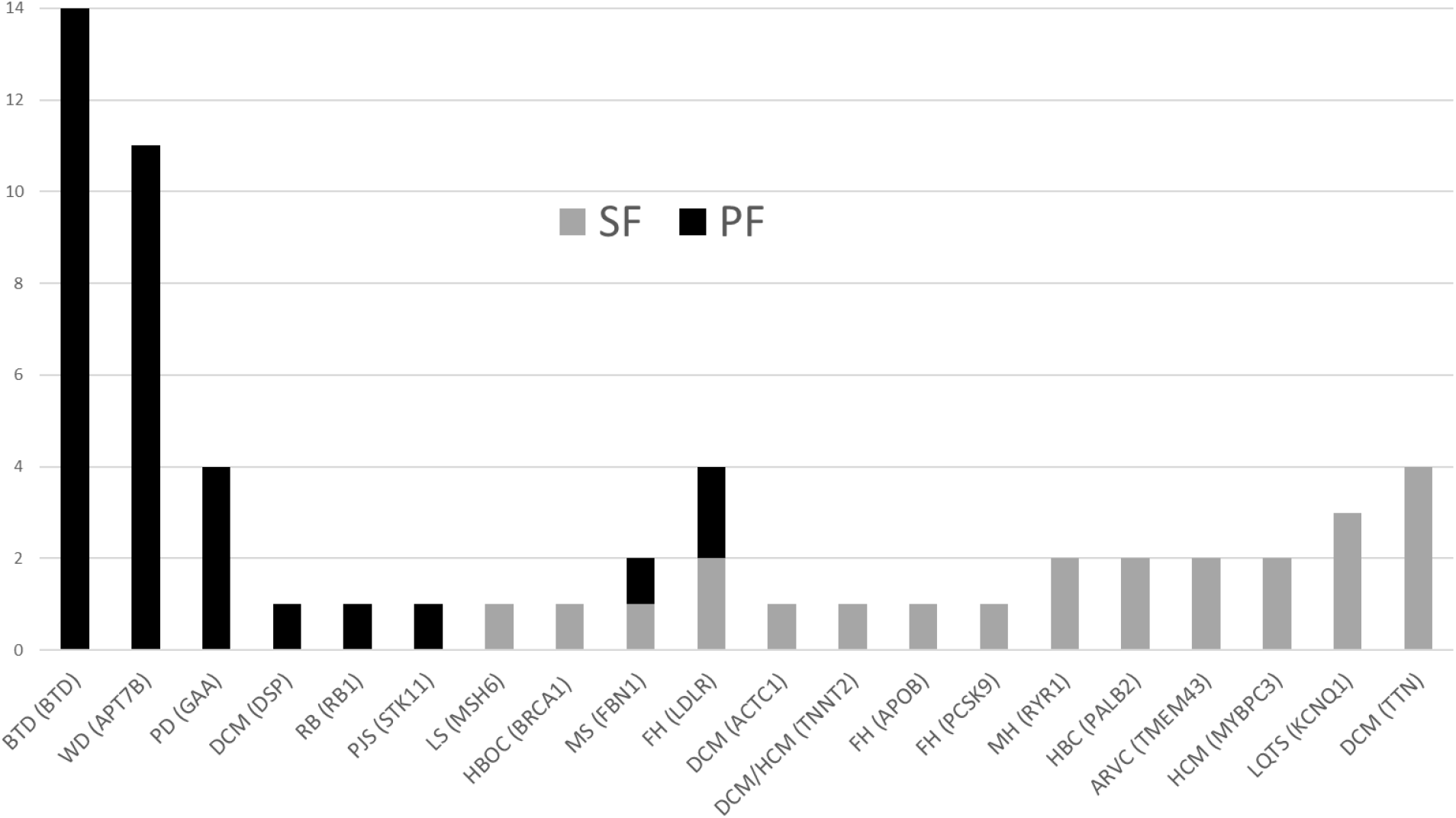
The ACMG secondary findings (SF) and ACMG primary findings (PF), diseases (genes). BTD - biotinidase deficiency; WD - Wilson disease; Pompe disease; DCM - dilated cardiomyopathy; RB - retinoblastoma; PJS - Peutz-Jeghers syndrome; LS - Lynch syndrome; HBOC - hereditary breast and ovarian cancer; MS - Marfan syndrome; FH - familial hypercholesterolemia; HCM - hypertrophic cardiomyopathy; MH - malignant hyperthermia; HBC - hereditary breast cancer; ARVC - arrhythmogenic right ventricular cardiomyopathy; LQTS - long QT syndrome.

#### Primary findings of non-ACMG secondary findings list (non-ACMG PF)

The non-ACMG PF were detected in 11/741 (1.5 %) symptomatic participants. For comparison to non-ACGM SF, we excluded VUS. The non-ACMG PF that included only P/LP variants were detected in 10/741 (1.3%) individuals. Compound heterozygous variants LP/VUS in *ABCA4* gene were found in one participant. We included this participant in further analysis, but *ABCA4* VUS was excluded from variant analysis. We found only biallelic (homozygous and compound heterozygous) mutations among the non-ACMG PF (100%). Totally, there were 11 unique P/LP variants in 3 genes related to 3 genetic diseases: *COL7A1* AR Epidermolysis bullosa dystrophica, *ABCA4* Stargardt disease type 1 and *CFTR* Cystic fibrosis (Supplement Table S2). Only one gene-disease pair occurred in both non-ACMG SF and PF groups: *ABCA4* Stargardt disease type 1. No identical variants were found in both groups.

## DISCUSSION

In our study, we aimed to define the frequency of secondary genomic findings based on the ACMG v3.0 (2021) gene list and the distribution of the associated disease type in the Pakistani population. Furthermore, based on criteria described in methods, we propose the inclusion of addition genes the SF list. Considering ACMG gene-disease pairs, our results demonstrated a prevalence of 2.7% SF in the Pakistani population.

The rates of ACMG SF in previous studies ranged from 0.6% (Jain et al. 2018) to 6.6% (Jang et al. 2015).

In the study of Aloraini T., *et al*., (Aloraini et al. 2021) the overall rate of SF in the Saudi population was higher than 8%. However, the rate of SF findings in 48 individuals (family members with the same SF variants were excluded) was calculated for the number of families (574) participated in the study. Probably the calculation of all participants with SF on the total number of participants (1254) would give a smaller rate.

The number of participants included in the SF studies ranged from less than two hundreds, 161 (Kuo et al. 2020), 196 (Jang et al. 2015), or less than three hundreds, 280 (Jalkh et al. 2020), to thousands, 6,240 (Elfatih et al. 2021), 21,915 (Gordon 2020). The studies with very large sample size can present some hurdles, such as ethical or financial, however, too small sample sized studies may prevent the findings from being extrapolated to population level (Faber and Fonseca 2014). Among the published studies, the highest frequencies of SF, 6.6% and 6.1%, were shown in the studies with relatively low numbers of participants, 196 and 280, respectively (Jang et al. 2015; Jalkh et al. 2020). Most of the SF studies represented local populations including Saudi Arabia (Aloraini et al. 2021), China (Chen et al. 2018), Qatar (Jain et al. 2018; Elfatih et al. 2021), Lebanon (Jalkh et al. 2020), Korea (Jang et al. 2015), Taiwan (Kuo et al. 2020), Netherlands (Haer-Wigman et al. 2019), Singapore (Jamuar et al. 2016), Thailand (Chetruengchai and Shotelersuk 2021). There were more representative studies: with self-reported race/ethnic groups including Hispanic or Latinx, Black or African American, Asian, American Indian, Alaska Native, or Pacific Islander, and White (Gordon 2020); East Asian ancestry (China, Vietnam) (Tang et al. 2018), European and African Americans (Natarajan et al. 2016); 1000 Genomes project, which includes 14 different populations in 4 major ancestry groups (Europe, East Asia, Africa, and the Americas) (Olfson et al. 2015). So far, to the best to our knowledge, no studies representing the Pakistani population and its ethnic groups have been conducted.

Depending on the time of publication, the studies used the v2.0 update of ACMG gene list, 2016 (59 genes) (Chen et al. 2018; Jain et al. 2018; Tang et al. 2018; Haer-Wigman et al. 2019; Gordon 2020; Jalkh et al. 2020; Kuo et al. 2020; Aloraini et al. 2021; Elfatih et al. 2021) or ACMG gene list v1.0, 2013 (56 genes) (Jang et al. 2015; Olfson et al. 2015; Jamuar et al. 2016; Natarajan et al. 2016; Hart et al. 2019). To our knowledge only one study was conducted to determine the frequency of P/LP variants in the 73 genes of ACMG v3.0 SF in which a SF frequency of 5.5% was reported (Chetruengchai and Shotelersuk 2022). Studies using ACMG v3.1 list have not yet been conducted. Considering the small difference in the number of genes between ACMG lists v1.0 and v2.0 (56 vs 59), a significant increase in SF was not expected. However, since 14 additional genes were added to the ACMG v3.0 and 5 additional genes were added to the ACMG 3.1 SF gene list, we addressed whether this upgrade would affect the frequency and change the disease structure of SF in future studies. In our study, the participants with a variant in *TTN* gene, exclusively present in v3.0 and v3.1 SF, resulting in an increase of 12.5%: 2.7% v3.1 vs 2.4% v2.0 ACMG SF lists.

Disease-causing copy number variants (CNV) account for 5 to 9% of hereditary diseases (Lindy et al. 2018; Truty et al. 2019; Brandt et al. 2020). Although CNV can be identified by whole exome sequencing (WES), the specificity and sensitivity limit their detection to larger alteration affecting at least three consecutive coding targeted exons. Most SF studies used exclusively WES (Jang et al. 2015; Natarajan et al. 2016; Chen et al. 2018; Haer-Wigman et al. 2019; Kuo et al. 2020; Aloraini et al. 2021) or both WES and WGS (Olfson et al. 2015; Jamuar et al. 2016; Jain et al. 2018; Hart et al. 2019). Only few SF studies used exclusively WGS for testing all individuals (Tang et al. 2018; Elfatih et al. 2021). Studies that used WGS can demonstrate a higher rate of SFs compared to those that used WES. For example, the substantial difference in the results of the two studies from the same country, Qatar, is probably in part due to the difference in the sequencing methods used. A predominantly WES (in 917 out of 1005 participants, 97%) was used in the first study published in 2018 with the SF rate of 0.6% (Jain et al. 2018). In the second study, 2021, WGS alone performed in all 6045 participants showed 2.3% of v2.0 ACMG SF (Elfatih et al. 2021). In fact, CNV SF variants would have been missed with WES.

Only one study reported the proportion of consanguinity rate (60%) among its participants (Aloraini et al. 2021). In our study, 79.9% participants were known to have consanguineous parents. In most studies SF were found as monoallelic mutations. Aloraini et al. in their study with 60% of consanguinity rate reported 1/49 (2.0%) LP variant biallelic (homozygous) in gene-disease pair with autosomal recessive (AR) mode of inheritance (Aloraini et al. 2021). In our study, 1/24 (4.2%) homozygous pathogenic variant (in *LDLR* gene) was found among ACMG SF and 8/16 (50.0%) biallelic (homozygous and compound heterozygous) mutations were found among non-ACMG SF. The studies on SF included symptomatic individuals for whom sequencing was performed for diagnostic purposes (Natarajan et al. 2016; Tang et al. 2018; Jalkh et al. 2020; Kuo et al. 2020), or both symptomatic and asymptomatic individuals (Jang et al. 2015; Jamuar et al. 2016; Chen et al. 2018; Gordon 2020; Aloraini et al. 2021; Elfatih et al. 2021). Two studies included exclusively healthy/asymptomatic participants (Jain et al. 2018; Haer-Wigman et al. 2019). The studies with only symptomatic individuals reported the rate of SF ranged from 1.0% (Natarajan et al. 2016) to 6.1% (Jalkh et al. 2020). The studies with only asymptomatic individuals reported the rate of SF ranged from 0.6% to 2.7% (Jain et al. 2018; Haer-Wigman et al. 2019). One study compared SF frequency among symptomatic and asymptomatic individuals, showing no difference: 7% for the healthy subjects (7/100) and 6% for the patients with a disease (6/96) (Jang et al. 2015). In our study, the rates of SF among symptomatic were higher, though not significantly, for both ACMG SF (3.0% vs 0.8%), and non-ACMG SF (2.0% vs 0.8%).

In previous SF studies PF were excluded from further analysis. In our study we further analysed PF and compare them to SF.

In contrast to the SF, biallelic variants predominated among the PF: 88.6% in ACMG PF and 100% in non-ACMG PF. Most gene-disease pairs were either only SF or only PF in both groups. Only two gene-disease pairs occurred in both groups, ACMG SF and PF: *FBN1* Marfan syndrome, and *LDLR* Familial hypercholesterolemia type 1. (Figure 3). No identical variants in these genes have been found in both groups. Only one gene-disease pair occurred in both non-ACMG SF and PF groups: *ABCA4* Stargardt disease type 1. No identical variants were found in both groups. We can conclude that diseases presenting as PF only (e.g., Wilson disease, biotinidase deficiency, Gaucher disease) are more clinically severe with earlier onset of manifestations. While other diseases detected as SF only (e.g., *TTN* and *ACTC1* related DCM) are less clinically severe and have later manifestation onset. The absence of the identical variants in *FBN1, LDLR, ABCA4* genes associated simultaneously with both SF and PF may indicate that the clinical severity and time of onset of these diseases may depend on characteristics of the variants.

In previous ACMG SF studies, cardiovascular diseases (CVD) and cancer predisposition conditions (CPC) were the leading disease types. The proportion of CVD ranged from 11.7% (Jalkh et al. 2020) to 54.5% (Haer-Wigman et al. 2019), the proportion of CPC ranged from 16.7% (Chen et al. 2018) to 46.2% (Jang et al. 2015). High percentage of CVD (75,0%) in our study, can be partly explained by different ACMG versions used. Using v3.0 we included 4 *TTN* variants. *TTN* cardiomyopathy is absent in previous versions of ACMG SF lists. This substantially increased the percentage of CVD and decreased the percentage of other conditions.

In the SF studies among CVD Brugada syndrome, LQTS and HCM were reported most frequently. BRCA1/2 hereditary breast and ovarian cancer (HBOC) and Lynch syndrome were most often reported cancer syndromes. Other frequently reported SF include familial hypercholesterolemia, malignant hyperthermia, Marfan syndrome, and Loeys-Dietz syndrome. (Jang et al. 2015; Olfson et al. 2015; Jamuar et al. 2016; Natarajan et al. 2016; Chen et al. 2018; Jain et al. 2018; Tang et al. 2018; Haer-Wigman et al. 2019; Hart et al. 2019; Gordon 2020; Jalkh et al. 2020; Kuo et al. 2020; Aloraini et al. 2021; Elfatih et al. 2021). In general, the structure of diseases in the SF mainly reflects the prevalence of diseases in the population. The diseases with high prevalence were detected with higher frequency: Brugada syndrome - 0.5 per 1000 (Vutthikraivit et al. 2018), HCM - 2 to 6 per 1000 (Batzner et al. 2019), HBOC - 2.9%-26.5% (Armstrong et al. 2019), Lynch syndrome - 0.44% (Haraldsdottir et al. 2017).

In our study *TTN* dilated cardiomyopathy type 1G; *KCNQ1* long QT syndrome type 1; *MYBPC3* hypertrophic cardiomyopathy type 4; *TMEM43* arrhythmogenic right ventricular dysplasia type 5 were the most frequent CVD; *MSH6* Lynch syndrome and *PALB2* hereditary breast cancer were the most frequent CPC among ACMG SF. (Figure 1). *ABCA4* Age-related macular degeneration type 2 was most frequent among non-ACMG SF. (Figure 2). We can assert that these diseases are frequent among the population of Pakistan. The overall prevalence of CAD in Pakistan ranges from 22 to 32% (Jafar et al. 2005). The prevalence of various cancers in Pakistan is high: the prevalence of breast cancer in females ranges from 20 to 50% and the prevalence of colorectal cancer in both males and females ranges from 4 to 6% (Idrees et al. 2018).

The studies reported individuals to be carriers of a disease allele in genetic disorders with AR mode of inheritance from ACMG SF list v2.0 with rates ranged from 0.40% (Jain et al. 2018; Elfatih et al. 2021) to 3.1% (Kuo et al. 2020). Despite the increase of number of AR diseases in ACMG v3.1 SF list, a smaller proportion of carriers (0.12%) was identified in out cohort. This could be due to low proportion of healthy individuals in our cohort compared to the studies mentioned above.

The first major **limitation** of our study is in the cohort, consisting of predominantly highly symptomatic young children. We would refrain from directly extrapolating the results obtained in this cohort to the population of Pakistan as a whole. The second major limitation is that the generation of the non-ACMG SF list is largely based on the opinion of experts familiar with the situation with genetic diseases in Pakistan.

## Conclusions

WGS is a reliable and easy test format to identify ACMG 3.1 and non-ACMG SF. The frequency of ACMG SF (2.7%) is within the range reported in most relevant studies. Higher proportion of CVD (75.0%) among ACMG SF in our study partly can be explained the additional inclusion of CVD (e.g., *TTN* DCM) in the ACMG v3.1 SF list. In addition, we reported a 1.9% rate of Pakistan specific non-ACMG SF and unexpectedly high proportion of biallelic variants among both ACMG SF (4.2%) and non-ACMG SF (50%). These results are relevant to epidemiology of Pakistan as a country with high rate of consanguineous marriages. Our findings may help guide the development of standards of practice in genomic medicine. As such, in countries with high levels of consanguinity, the ACMG criteria for SF can be expanded, and our list of non-ACMG SF is one example. Our findings also serve as a resource to inform decision-making in individuals undergoing genomic testing and drive future research.

## METHODS

### Participants

All participants were tested as part of clinical evaluation in Pakistani hospitals in Islamabad, Karachi, Lahore, Multan, and Peshawar for different reasons. Subjects were either index cases or healthy family members. The participants or guardians of the children participants signed the provided written informed consent forms. The study was approved by the Ethics Committee of Rostock University (Germany), A2022-0072, 25.04 2022.

### Whole genome sequencing and fata analysis

DNA samples were prepared using TruSeq DNA Nano Library Prep Kit from Illumina®. The libraries were pooled and sequenced with the 150 bp paired-end protocol on an Illumina platform to yield an average coverage depth of 30x for the nuclear genome. Raw read alignment to reference genome GRCH38 and variant calling, including single nucleotide substitutions (SNVs), small insertions/deletions (Indels) and structural variants (SVs) with default parameters were performed using DRAGEN (version 3.10.4, Illumina). SNV and indel annotation was performed by Varvis® (Limbus Medical Technologies GmbH; https://www.limbus-medtec.com/). Structural variants were annotated with ANNOTSV3.1 and in-house-structural variant database to obtain occurrence frequencies. Genetic variants are described following the Human Genome Variation Society (HGVS) recommendations (www.hgvs.org).

### Variant evaluation and interpretation

Only good quality variants with a minimum of 9 reads and an alternalive allele frequency of at least 0.3% were considered. Candidate variants were evaluated with respect to their pathogenicity and causality and categorized following ACMG guidelines (Richards et al. 2015) using the 5-tier classes: pathogenic, likely pathogenic, variants of uncertain significance (VUS), likely benign and benign. Variants were assessed in a routine diagnostic setting. An initial analysis was restricted to genes that have a clear association with patient’s phenotype using Human Phenotype Ontology nomenclature (HPO; https://hpo.jax.org/app/). During this step, common standards were followed. Briefly, the following aspects were considered: the minor frequency of the allele in control databases (gnomAD) and internal database, the in silico pathogenicity prediction and potential impact on respective protein, the known mechanism of the variant type-disease (missense, truncating etc.), segregation in the family, available external evidence OMIM (https://www.omim.org/), ClinVar information (https://www.ncbi.nlm.nih.gov/clinvar/), MasterMind (https://mastermind.genomenon.com/), and genotype-phenotype correlation.

Secondary findings that do not correlate with the provided phenotype(s) were reported according to ACMG recommendations for reporting of incidental findings in clinical exome and genome sequencing (Miller et al. 2021b). For this approach ACMG and non-ACMG genes were interrogated and patient’ phenotype was not considered. Only pathogenic and likely pathogenic variants with appropriate zygosity according to the known mode of inheritance for the corresponding gene (i.e., heterozygous for AD and homozygous or compound heterozygous for AR) were considered for reporting. Finally, the data was evaluated by screening the carrier status for both ACMG and non-ACMG-SF genes, including only heterozygous genetic variants, classified as pathogenic or likely pathogenic for AR diseases.

### The list of non-ACMG SF gene-phenotype pairs in Pakistani cohort

In addition to the ACMG SF, we have analysed a cohort of about 1000 Pakistani participants and identified gene-disease pairs with strong medical actionability, not yet included in the ACMG v3.1 secondary finding list. Based on this, we generated a list of Pakistani non-ACMG secondary findings (non-ACMD SF). (Supplement Table S3). Several aspects for inclusion were considered to generate the list of non-ACMG SF gene-phenotype pairs in Pakistani cohort. (1) Actionability: gene-phenotype pairs that scored a total of 10 or higher for actionability by the ClinGen Actionability Working Group (https://clinicalgenome.org/), or gene-phenotype pairs that scored a total of 10 or higher for actionability by expert opinion; (a) severity of disease; (b) penetrance or likelihood of disease development; (c) impact and/or burden of available treatment modalities or screening recommendations. (2) Regarding poor service for genetic diseases in Pakistan we did not exclude a) disorders that would typically be diagnosed clinically as well as b) disorders where a lifestyle, including diet, change is the prominent intervention. (3) Disorders where timing of the diagnosis was not critical for treatment efficacy were not included. (4) New-born screening (NBS) assays (e.g., Recommended Uniform Screening Panel, RUSP) are generally cheaper, more available, and sensitive to identify inborn error of metabolism than WGS. However, given the Pakistani specifics of NBS service, we did not exclude these conditions from the list. (5) Pharmacogenomic (PGx) findings which can be used to guide drug dose and choice can be potentially included.

## Data Availability

All data produced in the present work are contained in the manuscript.
All variants listed in the manuscript have been submitted to the NCBI ClinVar database (http://www.clinvar.com/) under submission ID: SUB11892244 and organization ID: 508679.

## DATA ACCESS

All variants listed in the manuscript have been submitted to the NCBI ClinVar database (http://www.clinvar.com/) under submission ID - SUB11892244 and organization ID – 508679.

## COMPETING INTEREST STATEMENT

AS, AR, GO, NA, VS declared employment in genomic testing company; HC, MH, NR, KR, RK did not declare any competing interest related to this study.

## ACKNOWLEDGMENTS

We thank the contributions of the administration and staff of The Children Hospital (Lahore), Pakistan Institute of Medical Sciences (Islamabad), Children Hospital and ICH (Multan), Town Women and Children Hospital (Peshawar), Liaquat National Hospital (Karachi), Arcensus laboratory and bioinformatics teams.

## AUTHOR CONTRIBUTIONS

AR, AS: design of the work; AR, VS, HC, MH, NR, KR, RK: acquisition of the data; GO, AR, AS, NA, VS: analysis, interpretation of the data, drafting the manuscript; AS: the lead in writing the manuscript; all authors provided critical feedback and helped shape the final version of the manuscript.

## Notes

### Competing Interest Statement

Aliaksandr Skrahin, Gabriela Oprea, Najim Ameziane, Arndt Rolfs, Volha Skrahina work in Arcensus genetic diagnostics company.

### Funding Statement

This study was funded by Arcensus GmbH
Goethe st. 20, 18055 Rostock,
phone: +49 171 4710484
Homepage: www.arcensus-diagnostics.com

### Author Declarations

Ethical approval of the study was granted by Universitat Rostock, Ethikkommission an der MedizinischenFakultat der Universitat Rostock / Ethics Committee of the Medical Faculty of Rostock University. St.-Georg-Str. 108, 18055 Rostock, Germany;; phone: +49 381 494 9939; homepage: www.ethik.med.uni-rostock.de

