## Supplementary material for "Secondary findings in a large Pakistani cohort tested with whole genome sequencing": SUplement Table S3

Supplement Table S3. Non-ACMG secondary finding list.

| Gene | Disease | MIM | MoI | Actionability | Disease type | Severity* | Likehood of disease** Level of evidence^ | Effectiveness of intervention*** Level of evidence^ | Nature of intervention **** | Total score |
|---|---|---|---|---|---|---|---|---|---|---|
| ABCA4 | Stargardt disease 1 | 248200 | AR | Expert | Eye disease | 2 | 3N | 2B | 3 | 10NB |
| ABCA4 | Age-related macular degeneration 2 | 153800 | AD | Expert | Eye disease | 2 | 3N | 2C | 3 | 10NC |
| ABCC6 | Pseudoxanthoma elasticum | 264800 | AR | Expert | Connective tissue | 2 | 3N | 2D | 3 | 10ND |
| ABCD1 | Adrenoleukodystrophy | 300100 | XLR | clinicalgenome.org | Metabolic | 2 | 3C | 3C | 3 | 11CC |
| ABCG8 | Sitosterolemia 1 | 210250 | AR | Expert | Metabolic | 2 | 3N | 3C | 2 | 10NC |
| AGXT | Primary hyperoxaluria 1 | 259900 | AR | Expert | Metabolic | 2 | 2N | 3A | 3 | 10NA |
| BAP1 | BAP1 Tumor predisposition syndrome | 614327 | AD | clinicalgenome.org | Cancer | 2 | 2C | 3D | 3 | 10CD |
| CFTR | Cystic fibrosis | 219700 | AR | eMERGE / Expert | Miscellaneous | 2 | 3A | 2A | 3 | 10AA |
| CLCN1 | Dominant myotonia congenita | 160800 | AD | Expert | Neuromuscular | 2 | 3D | 2A | 3 | 10DA |
| CLCN1 | Recessive myotonia congenita | 255700 | AR | Expert | Neuromuscular | 2 | 3D | 2A | 3 | 10DA |
| COL7A1 | Epidermolysis bullosa dystrophica | 131750 | AD | Expert | Skin disesae | 2 | 3B | 2D | 3 | 10BD |
| COQ8B | Nephrotic syndrome 9 | 615573 | AR | Expert | Urinary system | 3 | 2N | 2A | 3 | 10NA |
| DNAH5 | Primary ciliary dyskinesia 3 | 608644 | AR | Expert | Lung disease | 2 | 3N | 2A | 3 | 10NA |
| DUOXA2 | Thyroid dyshormonogenesis 5 | 274900 | AR | Expert | Endocrine | 2 | 3N | 2A | 3 | 10NA |
| EPCAM | Hereditary nonpolyposis colorectal cancer 8 (Lynch syndrome) | 613244 | AD | clinicalgenome.org | Cancer | 2 | 3A | 3A | 2 | 10AA |
| FCN3 | Immunodeficiency due to ficolin-3 deficiency | 613860 | AR | Expert | Immunodeficiency | 2 | 3N | 2D | 3 | 10ND |
| GCH1 | DOPA-responsive dystonia AD | 128230 | AD | clinicalgenome.org | Miscellaneous | 2 | 3C | 3C | 3 | 11CC |
| GCH1 | DOPA-responsive dystonia AR | 128230 | AR | clinicalgenome.org | Miscellaneous | 2 | 3C | 3C | 3 | 11CC |
| HFE | HFE-related hemochromatosis | 235200 | AR | clinicalgenome.org | Miscellaneous | 2 | 2A | 3B | 3 | 10AB |
| HMBS | Acute intermittent porphyria | 176000 | AD | clinicalgenome.org | Miscellaneous | 2 | 2C | 3B | 3 | 10CB |
| HNF1B | Maturity-onset diabetes of the young 5 | 125853 | AD | Expert | Endocrine | 2 | 3B | 3A | 3 | 11BA |
| IMPG1 | Vitelliform macular dystrophy 4 | 616151 | AR | Expert | Neuromuscular | 2 | 3N | 2C | 3 | 10NC |
| ITGB4 | Junctional epidermolysis bullosa 5A | 619816 | AR | Expert | Miscellaneous | 2 | 3B | 2A | 3 | 10BA |
| LMNA | Dilated cardiomyopathy 1A | 115200 | AD | clinicalgenome.org | Cardiovascular | 2 | 3B | 2N | 3 | 10BN |
| MEFV | Familial Mediterranean fever AR | 249100 | AR | clinicalgenome.org | Miscellaneous | 2 | 3C | 3A | 2 | 10CA |
| MFAP5 | Familial thoracic aortic aneurysm 9 | 616166 | AD | Expert | Cardiovascular | 3 | 3N | 2A | 3 | 11NA |
| MYO1H | Congenital central hypoventilation syndrome 2 | 619482 | AR | Expert | Neuromuscular | 3 | 3N | 2B | 3 | 11NB |
| POLD1 | Susceptibility to colorectal cancer 10 | 612591 | AD | clinicalgenome.org | Cancer | 2 | 3B | 3B | 2 | 10BB |
| POLE | Susceptibility to colorectal cancer 12 | 615083 | AD | clinicalgenome.org | Cancer | 2 | 3B | 3B | 2 | 10BB |
| PRKAR1A | Carney complex 1 | 160980 | AD | clinicalgenome.org | Miscellaneous | 3 | 2C | 3C | 3 | 11CC |
| SDHA | Hereditary paraganglioma/ pheochromocytoma | 614165 | AD | clinicalgenome.org | Cancer | 2 | 3C | 3B | 3 | 11CB |
| SERPINA1 | Alpha-1-antitrypsin deficiency | 613490 | AR | clinicalgenome.org | Miscellaneous | 2 | 3C | 3A | 2 | 10CA |
| SLC34A1 | Fanconi renotubular syndrome 2 | 613388 | AR | Expert | Miscellaneous | 2 | 3N | 2B | 3 | 10NB |
| SLC3A1 | Cystinuria | 220100 | AR | Expert | Urinary system | 2 | 3N | 2B | 3 | 10NB |
| SLC5A2 | Renal glucosuria | 233100 | AR | Expert | Urinary system | 2 | 3N | 2B | 3 | 10NB |
| SLC7A9 | Cystinuria | 220100 | AR | Expert | Urinary system | 2 | 3N | 2B | 3 | 10NB |
| TBC1D8B | Nephrotic syndrome 20 | 301028 | XL | Expert | Urinary system | 2 | 3N | 2A | 3 | 10NA |
| THBD | Atypical hemolytic uremic syndrome 6 | 612926 | AD | Expert | Hemolytic anemia | 3 | 2B | 3B | 2 | 10BB |
| VWF | von Willebrand disease 1 | 193400 | AD | clinicalgenome.org | Hemorrhagic | 2 | 3C | 3A | 2 | 10CA |

ACMG - the American College of Medical Genetics and Genomics; MIM - number from the 12th edition of Mendelian Inheritance in Man (MIM); MoI – mode of inheritance; *Severity: 3 - sudden death; 2 -possible death or major morbidity; 1 - modest morbidity; 0 - minimal or no morbidity; **Likehood of disease: 3 - >40% chance; 2 - 5-39% chance; 1 - 1-4% chance; 0 - <1% chance or unknown; ***Effectiveness of intervention: 3 - highly effective; 2 - moderately; 1 - minimally; 0 - ineffective / no intervention; ****Nature of intervention: 3 - low risk / medically acceptable / low intensity intervention; 2 - moderately acceptable / intensive intervention; 1 - greater risk / less acceptable / substantial intervention; 0 - high risk / poor acceptable / intensive / or no intervention; ^ Level of evidence: A - substantial evidence; B - moderate; C - minimal; D - poor; N - non-systematically or expert contributed evidence.
