## Supplement Table S1 for "Secondary findings in a large Pakistani cohort tested with whole genome sequencing"

Supplement Table S1. Secondary findings.

| Pat. no. | Gender | Consang. | Age, years | S/A | Gene | Disease | MIM | MOI | Disease type | Zygosity | Transcript ID | cDNA | Protein | Class. | Mut. | IoP | SF category |
|---|---|---|---|---|---|---|---|---|---|---|---|---|---|---|---|---|---|
| 1 | Female | Yes | 11-15 | S | *ACTC1* | Dilated cardiomyopathy 1R | 102540 | AD | Cardiovascular | Het | NM_005159.5 | c.843del | p.(Tyr281fs) | P | Deletion | In frame | ACMG |
| 2 | Female | Yes | 0-5 | S | *TNNT2* | Dilated cardiomyopathy 1D; hypertrophic cardiomyopathy 2 | 601494; 115195 | AD | Cardiovascular | Het | NM_001276345.2 | c.68-2A>G | p.? | LP | Substitution | Splicing mu | ACMG |
| 3 | Female | Yes | 6-10 | S | *FBN1* | Marfan syndrome | 154700 | AD | Cardiovascular | Het | NM_000138.5 | c.2563C>T | p.(Gln855*) | LP | Substitution | Nonsense | ACMG |
| 4 | Male | Yes | 0-5 | S | *TTN* | Dilated cardiomyopathy 1G | 245200 | AD | Cardiovascular | Het | NM_001267550.2 | c.32452G>T | p.(Glu10818*) | LP | Substitution | Nonsense | ACMG |
| 5 | Male | No | 0-5 | S | *RYR1* | Malignant hyperthermia 1 | 145600 | AD | Miscellaneous | Het | NM_000540.3 | c.4019del | p.(Gly1340Alafs*58) | LP | Deletion | Frameshift | ACMG |
| 6 | Male | Yes | 6-10 | S | *KCNQ1* | Long QT syndrome 1 | 192500 | AD | Cardiovascular | Het | NM_000218.3 | c.364dup | p.(Cys122Leufs*163) | LP | Duplication | Frameshift | ACMG |
| 7 | Female | Yes | 6-10 | S | *LDLR* | Familial hypercholesterolemia 1 | 143890 | AD | Cardiovascular | Het | NM_000527.5 | c.691T>G | p.(Cys231Gly) | LP | Substitution | Missense | ACMG |
| 8 | Male | Yes | 0-5 | S | *KCNQ1* | Long QT syndrome 1 | 192500 | AD | Cardiovascular | Het | NM_000218.3 | c.364dup | p.(Cys122Leufs*163) | LP | Duplication | Frameshift | ACMG |
| 9 | Male | No | 6-10 | S | *MYBPC3* | Hypertrophic cardiomyopathy 4 | 115197 | AD | Cardiovascular | Het | NM_000256.3 | c.3759dup | p.(Arg1254Glnfs*12) | LP | Duplication | Frameshift | ACMG |
| 9 | Male | No | 6-10 | S | *TTN* | Dilated cardiomyopathy 1G | 604145 | AD | Cardiovascular | Het | NM_001267550.2 | c.60457_60460 | p.(Val20153Profs*9) | LP | Deletion | Frameshift | ACMG |
| 10 | Female | Yes | 6-10 | S | *PCSK9* | Familial Hypercholesterolemia 3 | 603776 | AD | Cardiovascular | Het | NM_174936.4 | c.1069C>T | p.(Arg357Cys) | LP | Substitution | Missense | ACMG |
| 11 | Male | No | 6-10 | S | *TMEM43* | Arrhythmogenic right ventricular cardiomyopathy | 604400 | AD | Cardiovascular | Het | NM_024334.3 | c.393-2A>G | p.? | LP | Substitution | Splicing mu | ACMG |
| 12 | Male | No | 0-5 | S | *TMEM43* | Arrhythmogenic right ventricular cardiomyopathy | 604400 | AD | Cardiovascular | Het | NM_024334.3 | c.393-2A>G | p.? | LP | Substitution | Splicing mu | ACMG |
| 13 | Male | Yes | 0-5 | S | *KCNQ1* | Long QT syndrome 1 | 192500 | AD | Cardiovascular | Het | NM_000218.3 | c.1686-2A>G | p.? | LP | Substitution | Splicing mu | ACMG |
| 14 | Male | Yes | 0-5 | S | *MYBPC3* | Hypertrophic cardiomyopathy 4 | 115197 | AD | Cardiovascular | Het | NM_000256.3 | c.711C>A | p.(Tyr237*) | LP | Substitution | Nonsense | ACMG |
| 15 | Male | No | 0-5 | S | *MSH6* | Hereditary nonpolyposis colorectal cancer 5 | 614350 | AD | Cancer | Het | NM_000179.3 | c.2759del | p.(Lys920Argfs*25) | LP | Deletion | Frameshift | ACMG |
| 16 | Male | No | 31-35 | A | *RYR1* | Malignant hyperthermia 1 | 145600 | AD | Miscellaneous | Het | NM_000540.3 | c.14970-1G>A | p.? | LP | Substitution | Splicing mu | ACMG |
| 17 | Male | Yes | 0-5 | S | *APOB* | Familial hypercholesterolemia 2 | 144010 | AD | Cardiovascular | Het | NM_000384.3 | c.12578dup | p.(Ile4194Hisfs*2) | LP | Duplication | Frameshift | ACMG |
| 18 | Male | No | 0-5 | S | *BRCA1* | Hereditary breast and ovarian cancer | 604370 | AD | Cancer | Het | NM_007294.4 | c.3770_3771de | p.(Glu1257Glyfs*9) | P | Delins | Nonsense | ACMG |
| 19 | Female | Yes | 11-15 | S | *TTN* | Dilated cardiomyopathy 1G | 604145 | AD | Cardiovascular | Het | NM_001267550.2 | c.77493G>A | p.(Trp25831*) | LP | Substitution | Nonsense | ACMG |
| 20 | Female | Yes | 6-10 | S | *TTN* | Dilated cardiomyopathy 1G | 604145 | AD | Cardiovascular | Het | NM_001267550.2 | c.57545-2A>G | p.? | P | Substitution | Splicing mu | ACMG |
| 21 | Male | Yes | 0-5 | S | *LDLR* | Familial hypercholesterolemia 1 | 143890 | AR | Cardiovascular | Hom | NM_000527.5 | c.1897C>T | p.(Arg633Cys) | P | Substitution | Missense | ACMG |
| 22 | Male | Yes | 0-5 | S | *PALB2* | Hereditary breast cancer | 114480 | AD | Cancer | Het | NM_024675.4 | c.2229T>A | p.(Tyr743*) | LP | Substitution | Nonsense | ACMG |
| 23 | Male | Yes | 0-5 | S | *PALB2* | Hereditary breast cancer | 114480 | AD | Cancer | Het | NM_024675.4 | c.2353_2354de | p.? | LP | Deletion | Frameshift | ACMG |
| 24 | Female | Yes | 11-15 | S | *MUTYH* | MUTYH -associated polyposis | 608456 | AR | Cancer | Het | NM_001128425.2 | c.312C>A | p.(Tyr104*) | LP | Substitution | Nonsense | ACMG carrier |
| 25 | Female | Yes | 6-10 | S | *THBD* | Atypical hemolytic uremic syndrome 6 | 612926 | AD | Hemolytic anemia | Het | NM_000361.3 | c.245del | p.(Gly82Alafs*79) | LP | Deletion | Frameshift | non-ACMG |
| 26 | Male | Yes | 6-10 | S | *ABCA4* | Stargardt disease 1 | 248200 | AR | Eye | Hom | NM_000350.3 | c.6658C>T | p.(Gln2220*) | LP | Substitution | Nonsense | non-ACMG |
| 27 | Male | Yes | 0-5 | S | *ABCG8* | Sitosterolemia 1 | 210250 | AR | Metabolic | Hom | NM_022437.3 | c.2T>C | p.0? | LP | Substitution | Start loss | non-ACMG |
| 28 | Female | Yes | 0-5 | S | *ABCA4* | Age-related macular degeneration 2 | 153800 | AD | Eye | Het | NM_000350.3 | c.6658C>T | p.(Gln2220*) | LP | Substitution | Nonsense | non-ACMG |
| 29 | Female | Yes | 0-5 | S | *VWF* | von Willebrand disease 1 | 193400 | AD | Hemorrhagic | Het | NM_000552.5 | c.4222_4224de | p.(Lys1408del) | LP | Deletion | In frame | non-ACMG |
| 30 | Female | No | 0-5 | S | *SLC5A2* | Renal glucosuria | 233100 | AD | Urinary | Het | NM_003041.4 | c.885+5G>A | p.? | LP | Substitution | Splicing mu | non-ACMG |
| 31 | Male | Yes | 0-5 | S | *ABCA4* | Age-related macular degeneration 2 | 153800 | AD | Eye | Het | NM_000350.3 | c.214G>A | p.(Gly72Arg) | LP | Substitution | Missense | non-ACMG |
| 32 | Female | Yes | 0-5 | S | *COL7A1* | Epidermolysis bullosa dystrophica | 131750 | AD | Skin | Het | NM_000094.4 | c.2171-2A>G | p.? | LP | Substitution | Splicing mu | non-ACMG |
| 33 | Female | Yes | 0-5 | S | *COQ8B* | Nephrotic syndrome 9 | 615573 | AR | Urinary system | Hom | NM_024876.4 | c.1447G>T | p.(Glu483*) | LP | Substitution | Nonsense | non-ACMG |
| 34 | Male | Yes | 0-5 | S | *SLC34A1* | Fanconi renotubular syndrome 2 | 613388 | AR | Miscellaneous | Hom | NM_003052.5 | c.690dup | p.(Val231Serfs*32) | LP | Duplication | Frameshift | non-ACMG |
| 35 | Male | Yes | 0-5 | S | *DUOXA2* | Thyroid dyshormonogenesis 5 | 274900 | AR | Endocrine | Hom | NM_207581.4 | c.228G>C | p.(Trp76Cys) | LP | Substitution | Missense | non-ACMG |
| 36 | Male | Yes | 0-5 | S | *MFAP5* | Familial thoracic aortic aneurysm 9 | 616166 | AD | Cardiovascular | Het | NM_003480.4 | c.472C>T | p.(Arg158*) | P | Substitution | Nonsense | non-ACMG |
| 37 | Male | Yes | 51-55 | A | *IMPG1* | Vitelliform macular dystrophy 4 | 616151 | AD | Neuromuscular | Het | NM_001563.4 | c.1530del | p.(Gly511Valfs*9) | LP | Deletion | Frameshift | non-ACMG |
| 38 | Male | Yes | 0-5 | S | *MYO1H* | Congenital central hypoventilation syndrome 2 | 619482 | AR | Neuromuscular | Hom | NM_001101421.4 | c.2454+1G>A | p.? | LP | Substitution | Splicing mu | non-ACMG |
| 39 | Male | Yes | 0-5 | S | *AGXT* | Primary hyperoxaluria 1 | 259900 | AR | Metabolic | Comp het | NM_000030.3 | c.107G>A | p.(Arg36His) | LP | Substitution | Missense | non-ACMG |
| 39 | Male | Yes | 0-5 | S | *AGXT* | Primary hyperoxaluria 1 | 259900 | AR | Metabolic | Comp het | NM_000030.3 | c.32C>G | p.(Pro11Arg) | LP | Substitution | Missense | non-ACMG |
| 40 | Male | Yes | 16-20 | A | *DNAH5* | Primary ciliary dyskinesia 3 | 608644 | AR | Miscellaneous | Comp het | NM_001369.3 | c.12217G>A | p.(Val4073Met) | VUS | Substitution | Missense | non-ACMG |
| 40 | Male | Yes | 16-20 | A | *DNAH5* | Primary ciliary dyskinesia 3 | 608644 | AR | Miscellaneous | Comp het | NM_001369.3 | c.9514dup | p.(His3172Profs*15) | LP | Duplication | Frameshift | non-ACMG |
| 41 | Male | Yes | 36-40 | A | *CFTR* | Cystic fibrosis | 219700 | AR | Miscellaneous | Het | NM_000492.4 | c.1521_1523de | p.(Phe508del) | P | Deletion | In frame | non-ACMG carrier |
| 42 | Female | Yes | 36-40 | A | *CFTR* | Cystic fibrosis | 219700 | AR | Miscellaneous | Het | NM_000492.4 | c.1521_1523de | p.(Phe508del) | P | Deletion | In frame | non-ACMG carrier |
| 43 | Male | Yes | 36-40 | A | *CFTR* | Cystic fibrosis | 219700 | AR | Miscellaneous | Het | NM_000492.4 | c.1393-1G>A | p.? | P | Substitution | Splicing mu | non-ACMG carrier |
| 44 | Female | Yes | 36-40 | A | *CFTR* | Cystic fibrosis | 219700 | AR | Miscellaneous | Het | NM_000492.4 | c.1393-1G>A | p.? | P | Substitution | Splicing mu | non-ACMG carrier |
| 45 | Female | No | 6-10 | S | *CFTR* | Cystic fibrosis | 219700 | AR | Miscellaneous | Het | NM_000492.4 | c.1392G>T | p.(Lys464Asn) | LP | Substitution | Missense | non-ACMG carrier |
| 46 | Male | Yes | 0-5 | S | *ITGB4* | Junctional epidermolysis bullosa 5A | 619816 | AR | Miscellaneous | Het | NM_000213.5 | c.5218+2T>C | p.? | LP | Substitution | Splicing mu | non-ACMG carrier |
| 47 | Male | Yes | 0-5 | S | *MEFV* | AR familial Mediterranean fever | 134610 | AR | Miscellaneous | Het | NM_000243.3 | c.2177T>C | p.(Val726Ala) | P | Substitution | Missense | non-ACMG carrier |
| 48 | Female | Yes | 0-5 | S | *SLC7A9* | AR cystinuria | 220100 | AR | Urinary system | Het | NM_014270.5 | c.544G>A | p.(Ala182Thr | LP | Substitution | Missense | non-ACMG carrier |

Pat. – participant; Consang. – consanguinity; S/A – symptomatic/asymptomatic; MIM - number from the 12th edition of Mendelian Inheritance in Man (MIM); MoI – mode of inheritance; Class. – classification; Mut. – mutation type; IoP – impact on protein; PF – primary findings.
