## Supplement Table S2 for "Secondary findings in a large Pakistani cohort tested with whole genome sequencing"

Supplement Table S2. Primary findings.

| Pat. no. | Gender | Consang. | Age, years | Gene | Disease | MIM | MoI | Disease type | Zygosity | Transcript ID | cDNA | Protein | Class. | Mut. | IoP | PF |
|---|---|---|---|---|---|---|---|---|---|---|---|---|---|---|---|---|
| 1 | Male | Unknown | 6-10 | ATP7B | Wilson disease | 100066 | AR | Miscellaneous | Hom | NM_000053.4 | c.2930C>T | p.(Thr977Met) | P | Substitutio | Missense | ACMG |
| 2 | Female | Unknown | 6-10 | BTD | Biotinidase deficiency | 253260 | AR | Metabolic | Hom | NM_001370658.1 | c.40_41del | p.Gly14LeufsTer17 | P | Deletion | Frameshift | ACMG |
| 3 | Female | Unknown | 0-5 | BTD | Biotinidase deficiency | 253260 | AR | Metabolic | Hom | NM_001370658.1 | c.40_41del | p.Gly14LeufsTer17 | P | Deletion | Frameshift | ACMG |
| 4 | Male | Unknown | 0-5 | BTD | Biotinidase deficiency | 253260 | AR | Metabolic | Hom | NM_001370658.1 | c.41_44del | p.(Gly14Valfs*35) | LP | Deletion | Frameshift | ACMG |
| 5 | Male | Yes | 6-10 | ATP7B | Wilson disease | 277900 | AR | Miscellaneous | Hom | NM_000053.4 | c.3182G>A | p.(Gly1061Glu) | P | Substitutio | Missense | ACMG |
| 6 | Male | Yes | 6-10 | ATP7B | Wilson disease | 277900 | AR | Miscellaneous | Hom | NM_000053.4 | c.2513del | p.(Lys838SerfsTer35) | LP | Deletion | Frameshift | ACMG |
| 7 | Male | Yes | 0-5 | RYR1 | Malignant hyperthermia | 145600 | AD | Miscellaneous | Het | NM_000540.3 | c.2654G>A | p.(Arg885His) | VUS | Substitutio | Missense | ACMG |
| 8 | Female | Yes | 0-5 | BTD | Biotinidase deficiency | 253260 | AR | Metabolic | Hom | NM_001370658.1 | c.1386dup | p.(Glu463*) | LP | Duplicatior | Nonsense | ACMG |
| 9 | Male | Yes | 6-10 | BTD | Biotinidase deficiency | 253260 | AR | Metabolic | Hom | NM_001370658.1 | c.41_44del | p.(Gly14Valfs*35) | LP | Deletion | Frameshift | ACMG |
| 10 | Female | Yes | 0-5 | BTD | Biotinidase deficiency | 253260 | AR | Metabolic | Hom | NM_001370658.1 | c.41_44del | p.(Gly14Valfs*35) | LP | Deletion | Frameshift | ACMG |
| 11 | Female | Yes | 11-15 | ATP7B | Wilson disease | 277900 | AR | Miscellaneous | Hom | NM_000053.4 | c.2906G>A | p.(Arg969Gln) | P | Substitutio | Missense | ACMG |
| 12 | Female | Yes | 6-10 | ATP7B | Wilson disease | 277900 | AR | Miscellaneous | Hom | NM_000053.4 | c.2906G>A | p.Arg969Gln | P | Substitutio | Missense | ACMG |
| 13 | Male | Yes | 0-5 | BTD | Biotinidase deficiency | 253260 | AR | Metabolic | Hom | NM_001370658.1 | c.675C>A | p.(Cys225*) | P | Substitutio | Nonsense | ACMG |
| 14 | Male | Yes | 0-5 | BTD | Biotinidase deficiency | 253260 | AR | Metabolic | Hom | NM_001370658.1 | c.41_44del | p.(Gly14Valfs*35) | LP | Deletion | Frameshift | ACMG |
| 15 | Female | Yes | 0-5 | BTD | Biotinidase deficiency | 253260 | AR | Metabolic | Hom | NM_001370658.1 | c.1552C>A | p.(Arg518Ser) | P | Substitutio | Missense | ACMG |
| 16 | Female | No | 0-5 | GAA | Pompe disease | 232300 | AR | Metabolic; Cardiovascular | Hom | NM_000152.5 | c.1942G>A | p.(Gly648Ser) | P | Substitutio | Missense | ACMG |
| 17 | Male | Yes | 0-5 | BTD | Biotinidase deficiency | 253260 | AR | Metabolic | Hom | NM_001370658.1 | c.1552C>A | p.(Arg518Ser) | P | Substitutio | Missense | ACMG |
| 18 | Male | Yes | 0-5 | GAA | Pompe disease | 232300 | AR | Metabolic; Cardiovascular | Hom | NM_000152.5 | c.1930_1936dup | p.(Val646Glyfs*93) | LP | Duplicatior | Frameshift | ACMG |
| 19 | Female | No | 6-10 | BTD | Biotinidase deficiency | 253260 | AR | Metabolic | Hom | NM_001370658.1 | c.44_45del | p.(Cys15Leufs*16) | LP | Deletion | Frameshift | ACMG |
| 20 | Male | No | 0-5 | BTD | Biotinidase deficiency | 253260 | AR | Metabolic | Hom | NM_001370658.1 | c.44_45del | p.(Cys15Leufs*16) | LP | Deletion | Frameshift | ACMG |
| 21 | Male | Yes | 6-10 | ATP7B | Wilson disease | 277900 | AR | Miscellaneous | Hom | NM_000053.4 | c.3301G>A | p.(Gly1101Arg) | P | Substitutio | Missense | ACMG |
| 22 | Female | Yes | 6-10 | ATP7B | Wilson disease | 277900 | AR | Miscellaneous | Hom | NM_000053.4 | c.813C>A | p.(Cys271*) | P | Substitutio | Nonsense | ACMG |
| 23 | Female | Yes | 0-5 | BTD | Biotinidase deficiency | 253260 | AR | Metabolic | Hom | NM_001370658.1 | c.41_44del | p.(Gly14Valfs*35) | LP | Deletion | Frameshift | ACMG |
| 24 | Male | Yes | 6-10 | ATP7B | Wilson disease | 277900 | AR | Miscellaneous | Hom | NM_000053.4 | c.653C>T | p.(Ala218Val) | VUS | Substitutio | Missense | ACMG |
| 25 | Male | Yes | 26-30 | ATP7B | Wilson disease | 277900 | AR | Miscellaneous | Hom | NM_000053.4 | c.3147del | p.(Thr1050Hisfs*71) | P | Deletion | Frameshift | ACMG |
| 26 | Male | No | 6-10 | LDLR | Familial hypercholesterolemia 1 | 143890 | AD | Cardiovascular | Het | NM_000527.5 | c.2416dup | p.(Val806Glyfs*11) | P | Duplicatior | Frameshift | ACMG |
| 27 | Male | Yes | 11-15 | STK11 | Peutz-Jeghers syndrome | 175200 | AD | Cancer | Het | NM_000455.5 | c.640C>T | p.(Gln214*) | P | Substitutio | Nonsense | ACMG |
| 28 | Male | Yes | 11-15 | DSP | Dilated cardiomyopathy | 605676 | AR | Cardiovascular | Hom | NM_004415.4 | c.3959_3962del | p.(Ile1320Argfs*28) | LP | Deletion | Frameshift | ACMG |
| 29 | Female | No | 0-5 | GAA | Pompe disease | 232300 | AR | Metabolic; Cardiovascular | Hom | NM_000152.5 | c.1443G>A | p.(Trp481*) | P | Substitutio | Nonsense | ACMG |
| 30 | Male | Yes | 0-5 | GAA | Pompe disease | 232300 | AR | Metabolic; Cardiovascular | Hom | NM_000152.5 | c.925G>A | p.(Gly309Arg) | P | Substitutio | Missense | ACMG |
| 31 | Female | Yes | 11-15 | ATP7B | Wilson disease | 277900 | AR | Miscellaneous | Hom | NM_000053.4 | c.3182G>A | p.(Gly1061Glu) | P | Substitutio | Missense | ACMG |
| 32 | Female | No | 0-5 | RYR1 | Central core disease | 117000 | AD | Miscellaneous | Het | NM_000540.3 | c.10009C>T | p.(Arg3337Trp) | VUS | Substitutio | Missense | ACMG |
| 33 | Male | Yes | 6-10 | LDLR | Familial hypercholesterolemia 1 | 143890 | AR | Cardiovascular | Hom | NM_000527.5 | c.283T>C | p.(Cys95Arg) | P | Substitutio | Missense | ACMG |
| 34 | Female | No | 6-10 | FBN1 | Marfan syndrome | 154700 | AD | Cardiovascular | Het | NM_000138.5 | c.2243G>A | p.(Cys748Tyr) | P | Substitutio | Missense | ACMG |
| 35 | Female | Yes | 6-10 | TSC1 | Tuberous sclerosis 1 | 191100 | AD | Cancer | Het | NM_000368.5 | c.2025C>A | p.(Asp675Glu) | VUS | Substitutio | Missense | ACMG |
| 36 | Female | Yes | 0-5 | SDHB | Mitochondrial complex II deficiency nuclear 4 | 619224 | AR | Cancer | Hom | NM_003000.3 | c.304G>A | p.(Ala102Thr) | VUS | Substitutio | Missense | ACMG |
| 37 | Male | Yes | 11-15 | ATP7B | Wilson disease | 277900 | AR | Miscellaneous | Hom | NM_000053.4 | c.813C>A | p.(Cys271*) | P | Substitutio | Nonsense | ACMG |
| 38 | Male | Yes | 0-5 | RB1 | Retinoblastoma | 180200 | AD | Cancer | Het | NM_000321.3 | c.2663G>A | p.(Ser888Asn) | LP | Substitutio | Missense | ACMG |
| 39 | Male | No | 0-5 | BTD | Biotinidase deficiency | 253260 | AR | Metabolic | Comp het | NM_001370658.1 | c.41_44del | p.(Gly14Leufs*17) | LP | Deletion | Frameshift | ACMG |
| 39 | Male | No | 0-5 | BTD | Biotinidase deficiency | 253260 | AR | Metabolic | Comp het | NM_001370658.1 | c.44_45del | p.(Cys15Leufs*16) | LP | Deletion | Frameshift | ACMG |
| 40 | Male | No | 6-10 | TTN | Limb-girdle muscular dystrophy 10 | 608807 | AR | Cardiovascular | Comp het | NM_001267550.2 | c.39566_39568del | p.(Glu13189del) | VUS | Deletion | In frame | ACMG |
| 40 | Male | No | 6-10 | TTN | Limb-girdle muscular dystrophy 10 | 608807 | AR | Cardiovascular | Comp het | NM_001267550.2 | c.10770G>C | p.(Glu3590Asp) | VUS | Substitutio | Missense | ACMG |
| 41 | Female | No | 11-15 | ATP7B | Wilson disease | 277900 | AR | Miscellaneous | Comp het | NM_000053.4 | c.813C>A | p.(Cys271*) | P | Substitutio | Nonsense | ACMG |
| 41 | Female | No | 11-15 | ATP7B | Wilson disease | 277900 | AR | Miscellaneous | Comp het | NM_000053.4 | c.2998G>A | p.(Gly1000Arg) | LP | Substitutio | Missense | ACMG |
| 42 | Male | No | 0-5 | CFTR | Cystic fibrosis | 219700 | AR | Miscellaneous | Hom | NM_000492.4 | c.1753G>T | p.(Glu585*) | P | Substitutio | Nonsense | non-ACMG |
| 43 | Male | Yes | 0-5 | CFTR | Cystic fibrosis | 219700 | AR | Miscellaneous | Hom | NM_000492.4 | c.1705T>G | p.(Tyr569Asp) | LP | Substitutio | Missense | non-ACMG |
| 44 | Male | Yes | 0-5 | CFTR | Cystic fibrosis | 602421 | AR | Miscellaneous | Hom | NM_000492.4 | c.2856G>A | p.(Met952Ile) | LP | Substitutio | Missense | non-ACMG |
| 45 | Male | Yes | 0-5 | CFTR | Cystic fibrosis | 219700 | AR | Miscellaneous | Hom | NM_000492.4 | c.1753G>T | p.(Glu585*) | P | Substitutio | Nonsense | non-ACMG |
| 46 | Female | Yes | 0-5 | CFTR | Cystic fibrosis | 219700 | AR | Miscellaneous | Hom | NM_000492.4 | c.1393-1G>A | p.? | P | Substitutio | Splicing mu | non-ACMG |
| 47 | Male | Yes | 0-5 | CFTR | Cystic fibrosis | 219700 | AR | Miscellaneous | Hom | NM_000492.4 | c.1393-1G>A | p.? | P | Substitutio | Splicing mu | non-ACMG |
| 48 | Male | Yes | 11-15 | CLCN1 | AR Myotonia congenita | 255700 | AR | Neuromuscular | Hom | NM_000083.3 | c.1190T>A | p.(Val397Asp) | VUS | Substitutio | Missense | non-ACMG |
| 49 | Male | Yes | 0-5 | CFTR | Cystic fibrosis | 219700 | AR | Miscellaneous | Hom | NM_000492.4 | c.223C>T | p.(Arg75*) | LP | Substitutio | Nonsense | non-ACMG |
| 50 | Male | Yes | 0-5 | COL7A1 | AR Epidermolysis bullosa dystrophica | 226600 | AR | Skin | Hom | NM_000094.4 | c.5124+2T>C | p.? | LP | Substitutio | Splicing mu | non-ACMG |
| 51 | Female | Yes | 0-5 | ABCA4 | Stargardt disease 1 | 619157 | AR | Eye | Comp het | NM_000350.3 | c.5882G>A | p.(Gly1961Glu) | LP | Substitutio | Missense | non-ACMG |
| 51 | Female | Yes | 0-5 | ABCA4 | Stargardt disease 1 | 619157 | AR | Eye | Comp het | NM_000350.3 | c.5278C>A | p.(Leu1760Ile) | VUS | Substitutio | Missense | non-ACMG |
| 52 | Female | No | 0-5 | CFTR | Cystic fibrosis | 219700 | AR | Miscellaneous | Comp het | NM_000492.4 | c.1393-1G>A | p.? | P | Substitutio | Splicing mu | non-ACMG |
| 52 | Female | No | 0-5 | CFTR | Cystic fibrosis | 219700 | AR | Miscellaneous | Comp het | NM_000492.4 | c.2T>C | p.0? | P | Substitutio | Start loss | non-ACMG |

Pat. – participant; Consang. – consanguinity; MIM - number from the 12th edition of Mendelian Inheritance in Man (MIM); MoI – mode of inheritance; Class. – classification; Mut. – mutation type; IoP – impact on protein; PF – primary findings; Hom – homozygous; Het – heterozygous; comp het – compound heterozygous.
